# Spatially Aware Transformer Networks for Contextual Prediction of Diabetic Nephropathy Progression from Whole Slide Images

**DOI:** 10.1101/2023.02.20.23286044

**Authors:** Benjamin Shickel, Nicholas Lucarelli, Adish S. Rao, Donghwan Yun, Kyung Chul Moon, Seung Seok Han, Pinaki Sarder

## Abstract

Diabetic nephropathy (DN) in the context of type 2 diabetes is the leading cause of end-stage renal disease (ESRD) in the United States. DN is graded based on glomerular morphology and has a spatially heterogeneous presentation in kidney biopsies that complicates pathologists’ predictions of disease progression. Artificial intelligence and deep learning methods for pathology have shown promise for quantitative pathological evaluation and clinical trajectory estimation; but, they often fail to capture large-scale spatial anatomy and relationships found in whole slide images (WSIs). In this study, we present a transformer-based, multi-stage ESRD prediction framework built upon nonlinear dimensionality reduction, relative Euclidean pixel distance embeddings between every pair of observable glomeruli, and a corresponding spatial self-attention mechanism for a robust contextual representation. We developed a deep transformer network for encoding WSI and predicting future ESRD using a dataset of 56 kidney biopsy WSIs from DN patients at Seoul National University Hospital. Using a leave-one-out cross-validation scheme, our modified transformer framework outperformed RNNs, XGBoost, and logistic regression baseline models, and resulted in an area under the receiver operating characteristic curve (AUC) of 0.97 (95% CI: 0.90-1.00) for predicting two-year ESRD, compared with an AUC of 0.86 (95% CI: 0.66-0.99) without our relative distance embedding, and an AUC of 0.76 (95% CI: 0.59-0.92) without a denoising autoencoder module. While the variability and generalizability induced by smaller sample sizes are challenging, our distance-based embedding approach and overfitting mitigation techniques yielded results that suggest opportunities for future spatially aware WSI research using limited pathology datasets.

## 1. INTRODUCTION

Diabetic nephropathy (DN) in the setting of type 2 diabetes is one of the leading drivers of chronic kidney disease (CKD) and end-stage renal disease (ESRD) worldwide [1]. The incidence of DN is growing due to increasing prevalence of type 2 diabetes, where complications such as proteinuria and decline in kidney function affect more than 40% of patients [2-4]. Kidney biopsy is the gold standard for the diagnosis of kidney disease, and histology aids in predicting patient outcomes and response to therapy [5]. However, the heterogenous nature of DN, the addition of non-diabetic pathology, and the unclear association between kidney histology and clinical markers [6] present barriers to a comprehensive approach to developing personalized treatment plans based on disease trajectory and long-term outcomes [7]. While studies have developed ESRD prediction models of DN in type 2 diabetes in patients [3, 8-10], to our knowledge none have sufficiently integrated the complexities, including spatial organization at the whole slide level, of renal pathology images for patient-level clinical prediction.

The spatial distribution of renal pathology can indicate multidimensional kidney function and can be useful for patient-level prognostication and guiding therapies [11, 12], but quantifying the relative distances between compartments (which can appear in various stages of functional health) can be difficult or impossible for a pathologist to precisely determine in large whole slide images (WSI). In this work, we present an artificial intelligence (AI) approach for automatically generating patient-level ESRD predictions from the spatially aware morphological feature extraction of glomeruli present in whole slide periodic acid-Schiff (PAS) images using transformers, a recent class of deep learning (DL) model with increasing prevalence in natural language processing (NLP) and computer vision (CV) domains. While studies have shown the ability of machine learning (ML) to predict ESRD from clinical variables [13-15] and pathologist-interpreted quantification of histology images [16, 17] with varying accuracy, to our knowledge we are the first to develop an AI approach integrating handcrafted histology image feature extraction, nonlinear dimensionality reduction, pairwise distance-based relative positional encoding, and deep transformer networks for whole slide contextualization for patient-level ESRD prediction.

Recent advancements in digital pathology have been driven by technical breakthroughs in the field of AI known as DL, the subfield of ML encompassing models that have proven strikingly adept at extracting and learning from complex spatial information contained in raw pixels. Several studies have demonstrated the ability for DL to achieve pathologist-level performance for many common tasks such as structural segmentation from pathology images [9, 18-21] and diagnostic applications [18, 22-27]. The majority of AI-enabled digital pathology applications have been developed using the convolutional neural network (CNN). While the CNN has dominated the computer vision landscape of the past decade across both medical and non-medical domains, a new DL modeling paradigm has recently emerged known generally as the *transformer* [28].

Transformers originated from and are most publicized in the NLP domain (e.g., BERT [29], GPT-3 [30], T5 [31]), including notable examples in the medical domain [32-35], and have begun to demonstrate state-of-the-art performance in general imaging applications [36-38] including computational pathology [39-41]. While the original transformer used an encoder-decoder architecture due to its primary task of machine translation [28], many modern transformers for classification tasks (including ours) use an encoder-only architecture. Mechanistically, transformers are predominantly composed of stacked self-attention layers. The concept of attention in DL consists of methods for computing an alignment or similarity score between two vectorized representations, typically corresponding to a discrete input element such as a substring token or an image patch. Because transformers encode each input element as a linear combination of all other inputs, they are adept at capturing context that may be important for a given application. Because of the flexible nature of self-attention, transformers can be designed with a variety of inputs (including multimodal implementations [42]). In this paper, we develop a hybrid framework that capitalizes on both the advantages of established expert-defined morphological features, and the data-driven contextualization of sequential transformer models.

The past two years have seen a surge in popularity of transformer modeling for common computational pathology tasks such as WSI segmentation [40, 43, 44] and histology image classification [41, 45-47]. Transformers have also been used for pathologist-level question-answering from histological imaging [39], predicting pathologists’ visual attention [48], and for pathology text mining [49]. Nearly all applications of transformer-based approaches to whole slide imaging implement vision transformers (ViT), including recent works that combine CNNs and transformers [45]. In contrast, our multi-stage pipeline extracts expert-defined features from segmented structures before deriving WSI context using transformer encoders.

Attention mechanisms have historically been used in a variety of temporal DL models not only for improving performance through a more sophisticated representation of context, but also as an explainability tool for justifying model predictions by guiding internal focus to the temporal elements that have greatest impact on a particular prediction task. A post-hoc analysis of attention scores can provide human-interpretable explanations of how a model is representing a given patient or application. Because transformers are uniquely composed almost entirely of attention mechanisms, the opportunity for enhanced transparency can lead to better pathologist and clinical understanding of AI-driven predictions and facilitate a higher level of practitioner trust in prognostication and patient-centered decision making.

Furthermore, since temporality is represented in self-attention networks through the inclusion of an *a priori*-computed positional embedding rather than computationally time-consuming serial element processing, transformers are highly parallelizable and can be easily scaled up to distributed training environments. Our transformer framework also demonstrates the utility of self-attention networks in conjunction with careful selection of positional encodings to naturally represent variable-length sets of input elements (as opposed to a CNN or recurrent neural network [RNN], which assumes an implicit input ordering). In this paper, each WSI is represented by a slide-dependent number of glomeruli.

Our contributions can be summarized by the following:

- We introduce a multi-stage, hybrid AI framework integrating DL-based segmentation, handcrafted histological feature extraction, nonlinear feature compression, self-attention instance pooling, and deep transformer networks for modeling variable-length sets of WSI glomeruli and generating patient-level ESRD predictions more accurately than RNNs, XGBoost, or logistic regression models.
- We develop a graph-inspired formulation of relative positional encoding that integrates prior Euclidean distance measurements between every pair of glomeruli segmented from a WSI and show that it improves ESRD prediction AUC by up to 10%.
- We implement a collection of overfitting mitigation techniques and demonstrate that advanced DL methods can be used with small or restricted datasets with careful application.
- Compared with our prior work utilizing an RNN for WSI instance pooling, which fundamentally assumes an implicit temporal or spatial ordering, we demonstrate that self-attention networks present a more natural fit for contextualizing variable-length sets of renal structures present in WSIs that results in more accurate clinical predictions.
- We lay the groundwork for future hybrid transformer approaches for patient-level clinical forecasting from WSI data representations and multimodal augmentations with laboratory results and other clinical variables.

## 2. METHODOLOGY

We developed a novel multi-stage feature extraction and DL classification framework for predicting the onset of ESRD within two years from renal biopsy WSIs. Our pipeline consists of multiple stages that address challenges of applying DL to relatively small sample sizes by integrating handcrafted feature extraction with supervised ML (**Figure 1**). The prediction framework was designed to integrate spatial relationships among glomeruli based on recent literature [11, 12] that can improve the derivation of a whole slide context for WSI prediction for each patient biopsy.

**Figure 1.**
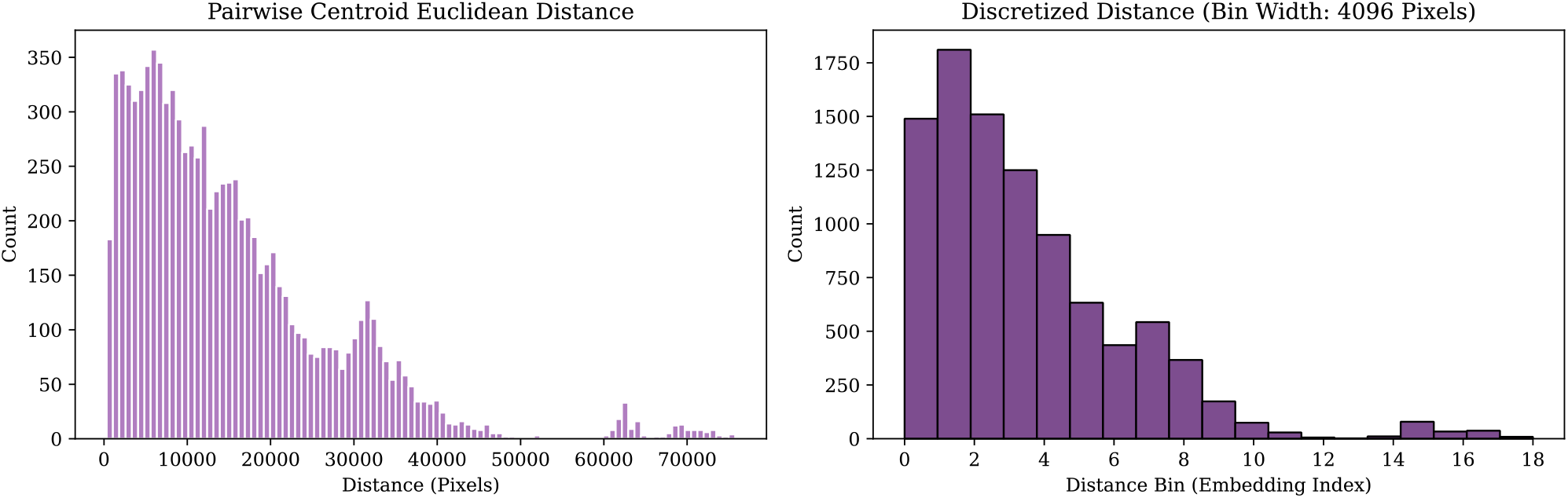
Distribution of pixel distances between the computed centroids of every pair of segmented glomeruli across all 56 WSIs (left) and corresponding integer discretization identifiers (right). The calculated distance between each pair of glomeruli is assigned an integer bin (bin width: 4,096 pixels) that is used to index an embedding lookup table for relative distance-based positional encoding in the modified self-attention process (Equation 3).

The goal of our framework is to generate accurate patient-level ESRD predictions from instance features extracted from a set of visible glomeruli contained in biopsy WSIs. This type of prediction setting can be viewed as a type of multiple-instance learning (MIL), a broad set of weakly supervised ML techniques with a long history in digital pathology for classifying individual or complete sets (“bags”) of image-based instances [50-52]. Among others, one of the central tasks of MIL is the development of bag embeddings based on a variable number of instances contained in each bag (e.g., in our dataset, each WSI contains a variable number of observable glomeruli). In this study, we demonstrate the effectiveness of transformers and self-attention [28] for this purpose, and develop a multi-stage pipeline that includes nonlinear, compressed representations of handcrafted features (as opposed to learning directly from raw pixels) that can alleviate the challenges of working with smaller histology datasets.

### 2.1 Histological Feature Extraction

The first stage consisted of a panoptic segmentation step to isolate the glomeruli contained in each patient WSI. The second stage involved the extraction of (1) expert-defined color, texture, and morphological features, (2) binary indication of sclerosis, and (3) the two-dimensional glomerulus centroid location from each segmented glomerulus. In this study, we utilize an existing dataset of 56 patient biopsies that we curated in prior work [53]. A more detailed description of data acquisition and processing are described in Section 3.

Following *a priori* data acquisition, segmentation, and handcrafted feature extraction (Section 3), each detected glomerulus was represented by a set of handcrafted features *x*_*glom*_ ∈ ℝ^1 × *d*^ denoting global sclerosis, texture, color, and morphology. In this study, we build upon our previous feature extraction pipeline [53], which generates *d* = 316 numerical features per glomerulus. We obtained a single WSI per patient, and since each WSI was associated with a variable number of observable glomeruli, each patient (i.e., each sample in our dataset) was represented by 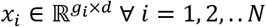, where *g*_*i*_ is the number of distinct glomeruli segmented from patient *i*’s WSI, and *N* is the number of patients in our dataset. In this study, we extracted features for *N* = 56 patients.

### 2.2 Denoising Autoencoders for Dimensionality Reduction

While the feature extraction process yielded a large and comprehensive set of glomerulus attributes, we empirically determined that not all were helpful in predicting a patient’s two-year ESRD status when passed to downstream ML models. We hypothesized that, given our limited dataset size (N = 56), the inclusion of our full suite of imaging features was resulting in problematic overfitting to potentially irrelevant noise instead of recognizing the true signal. Our final prediction framework includes a dimensionality reduction step designed to reduce the overall feature space and transform the imaging features into a more meaningful representation.

We developed a denoising autoencoder (DAE) [54] to derive a compressed representation of each glomerulus vector *x*_*glom*_, independent of the number of segmented glomeruli in each WSI. Briefly, the DAE is a nonlinear encoder-decoder DL model in which the encoder learns *h*_*i*_, a compressed and lower-dimensional hidden representation of an input *x*_*i*_, and the decoder learns to reconstruct the input *x*_*i*_ using the compressed transformation *h*_*i*_. The architecture bottleneck induced by the compression of the inner hidden layers results in a more information-dense and dimensionally reduced embedding of each input. A tunable proportion of masking noise (achieved here with dropout) is added to the input *x*_*i*_, which tends to improve the quality of input embeddings by forcing the model to reconstruct missing inputs based on a robust latent representation *h*_*i*_.

In this study, our DAE consists of an encoder with four fully connected layers that transform the original dimensionality of *d* = 316 to 128, 64, 32, and finally 16 latent features. Each fully connected layer is followed by rectified linear unit activation and a dropout layer. Our decoder is a complementary reversal of the encoder, with linear-activation-dropout blocks of 32, 64, 128, and *d* = 316 dimensions. No activation or dropout is added to the final decoder layer, and the DAE is trained to minimize mean squared reconstruction error. All DAEs were trained with an Adam optimizer with learning rate of 0.001, L2 regularization of 0.0001, and dropout rate of 20%.

Given our leave-one-out cross validation experimental design, we trained a separate DAE on each of the 56 unique training folds to prevent any potential data leakage from the test instance. After each autoencoder was trained, all *x*_*glom*_ feature vectors were transformed into lower-dimensional embeddings by passing them through the DAE encoder sub-network, resulting in transformed glomerulus representations of 16 dimensions (instead of the original 316) and overall initial WSI representations given by 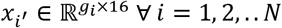. While the encoded feature representations are fixed after pre-training the DAE, our final transformer model includes an additional learnable fully connected layer to allow the model to further transform the latent glomeruli vectors for optimal ESRD prediction.

### 2.3 Contextualizing WSIs with Sequential Transformers and Self-Attention

Unlike many recent applications of transformers to pathology images [55], a traditional vision transformer (ViT) is not suitable for our framework because we employ an expert-defined feature extraction step and do not operate on raw pixel values. Furthermore, our model inputs are sparse (one feature vector per glomerulus), which differs from the high-resolution ViT approaches that can be prone to overfitting with a smaller dataset. Instead, we adapt the style of transformer that is typically used for natural language processing (NLP) applications. Specifically, we base our model on the architecture of Bidirectional Encoder Representations from Transformers (BERT) [56], a model typically used for representing text input; instead of a sequence of word token embeddings, we use a sequence of glomerulus features.

The basic flow of our prediction model is as follows. First, a set of all DAE-encoded glomeruli features from a patient’s WSI is passed into the DL model. A fully connected layer with Gaussian error unit activation and 20% dropout performs a further learnable transformation of each glomerulus vector into a hidden representation. The set of these instance representations is passed through a BERT transformer classifier, which introduces a special global classification token. While each glomerulus token learns to represent itself through a weighted sum of pairwise alignments with all other glomeruli through the self-attention process, the special classification token captures a more global context among all glomeruli. The global context vector is passed through a fully connected output layer to generate a probability of two-year ESRD.

### 2.4 Graph-Inspired Pairwise Distance Embeddings

Transformers are inherently temporally and spatially agnostic, at their core relying on simple self-attention mechanisms (capturing similarity or alignment) between every pair of input tokens. For applications where the order (e.g., words in natural language) or structure (e.g., pixels in computer vision) of inputs is important, a *positional embedding* is added to the representation of each input token before the self-attention process. Positional embeddings are typically based on absolute position of a token, such as the word number in a sentence (one-dimensional) or the (x, y) coordinates of a pixel or patch in an image (two-dimensional). However, in our application, the order of segmented glomeruli does not contain any inherent or natural order. Furthermore, the absolute (x, y) position of each glomerulus carries little meaning in a WSI, except for their relation to other glomeruli. With enough training data, a transformer can be expected to learn spatial relationships from enough examples of tokens equipped with absolute coordinates; but, given our limited dataset, learning such interactions from scratch is likely infeasible.

One significant contribution of our work is the adaptation of recent transformer literature on *relative positional embeddings* [57, 58] to spatial applications, and integration of data-driven spatial relationships with prior knowledge extracted from WSIs. Inspired by recent methods to induce graph structure within transformer architectures [59], we develop a pairwise relative positional embedding based on the Euclidean distance between every pair of glomeruli centroids (**Figure 1**). Instead of adding any independent spatial information to each token (e.g., embeddings based on raw positional coordinates), we inject relative distance-based embeddings during the pairwise self-attention step.

For each set *G*_*i*_ of observable glomeruli in patient *i*’s WSI (each patient’s image contains |*G*_*i*_ | glomeruli), we developed a pairwise distance matrix 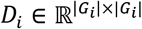 that computes the Euclidean distance between the centroids of every pair of glomeruli as measured in pixels:

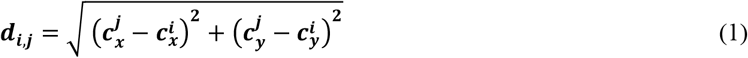

where 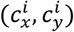 is the computed centroid of glomerulus *i*, and 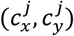 is the computed centroid of glomerulus *j* We modified the standard pairwise self-attention operation from Vaswani et al. [28] (Equation 2)

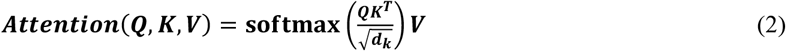

to add to the pairwise dot product calculations a learned embedding matrix 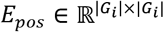 calculations, indexed by the relative pairwise centroid distance between every pair of glomeruli extracted from a given WSI (Equation 3):

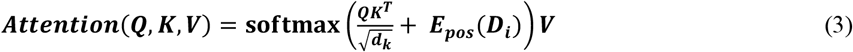

By replacing the standard self-attention equation (Equation 2) with the spatially aware modification (Equation 3) and parameterizing with prior Euclidean distances computed during the segmentation step, the transformer prediction model can learn to weight every pairwise glomeruli interaction by relative spatial proximity in addition to alignment of imaging characteristics. In this study, we discretized continuous Euclidean distances between glomeruli centroids to consecutively numbered bins of width 4096 pixels based on prior ViT work with cancer histology [55]. Our discretization process allows integer lookup into the *E*_*pos*_ embedding table and provides an additional measure of regularization that is important in our limited data setting. Our dataset contained 19 unique bins of discretized pairwise centroid distances.

### 2.5 Experiment Details

The goal of our system was to predict presence of ESRD two years after initial biopsy, using only imaging features and centroid distances from each patient’s WSI. Our dataset of renal biopsies is described in detail in Section 3. In this study, we compared our multi-stage transformer network (**Figure 2**) to several baseline approaches, including a previously established RNN architecture from our prior work predicting RPS classification of diabetic nephropathy [9]. We also experimented with XGBoost and logistic regression baseline models using a tabular dataset created by averaging all 316 histological features. All models were evaluated based on area under the receiver operating characteristic curve (AUROC), area under the precision-recall curve (AUPRC), accuracy, sensitivity, and specificity. For all metrics, 95% confidence intervals were generated using a bootstrapping procedure with 100 iterations.

**Figure 2.**
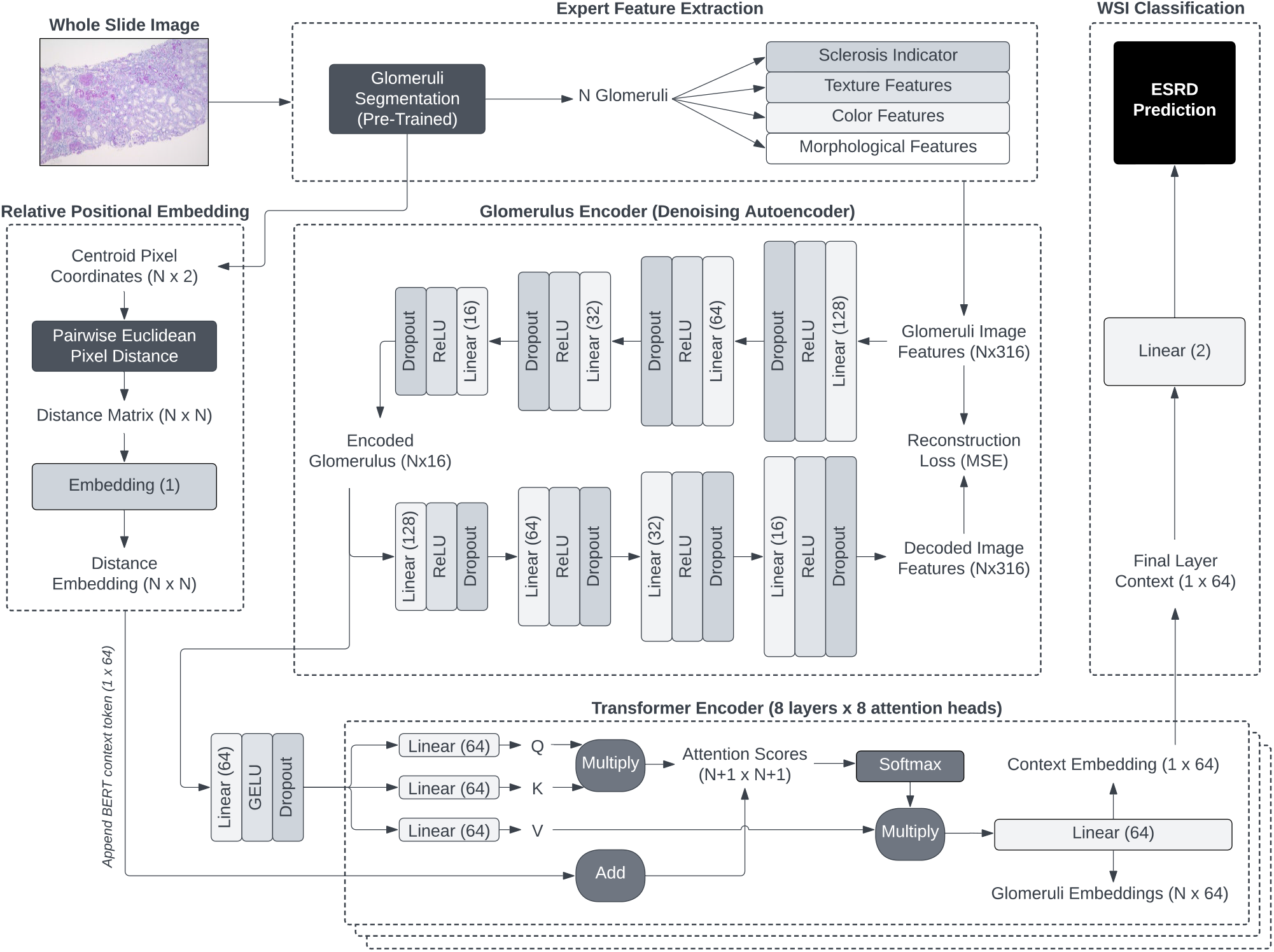
Multi-stage whole slide images (WSIs) processing pipeline, deep learning representation, and end-stage renal disease prediction framework. Glomeruli are first extracted from WSIs using a pretrained panoptic segmentation network. Next, handcrafted expert-defined features corresponding to color, texture, morphology, sclerosis, and centroid position are extracted from each glomerulus. A compressed feature representation is generated by training a denoising autoencoder and extracting the glomerulus encoding from the innermost hidden layer. Euclidean distances between every pair of glomeruli in a WSI are computed, discretized, and used as indices in an embedding lookup table for integration with transformer self-attention. The variable, slide-specific number of glomeruli feature vectors are contextualized through a deep self-attention transformer network, and finally aggregated into a static slide representation vector that is used to predict two-year end-stage renal disease.

To mitigate overfitting challenges applying DL using our relatively small sample size (N = 56) and to accurately estimate our framework’s generalizability, we report results using leave-one-out cross-validation. This process involves training 56 different models from a training set of 55 WSIs and generating a prediction on the remaining one WSI. This process is repeated with each WSI as the test sample until a prediction is generated for all 56 patients. During each of the 56 iterations, we normalized features, trained a separate DAE, and trained a separate transformer prediction network using only the 55 training samples.

The goal of this study is to demonstrate feasibility of a distance-based self-attention process for WSI contextualization and histological ESRD prediction. With that in mind, we did not perform an exhaustive hyperparameter search, instead selecting reasonable initial values and performing small deviation experiments to measure effects of a select number of important parameters (see **Table 1**). Hyperparameters for the DAE are described in detail in Section 2.2. The transformer module was composed of 8 layers with 8 attention heads per layer. All hidden layers in the prediction model used 64 hidden units with Gaussian Error Linear Unit (GELU) activations, including the feedforward component of the transformer encoder.

**Table 1.** Two-year end-stage renal disease prediction results using leave-one-out cross-validation. Model hyperparameters that deviate from baseline setting (D = 0.5, L2 = 1e-4, LR = 1e-2) are shown in parentheses. D: dropout, L2: L2 regularization, LR: learning rate, DAE: denoising autoencoder, RNN: recurrent neural network, Logistic: logistic regression, AUROC: area under the receiver operating characteristic curve, AUPRC: area under the precision-recall curve.

| Model | Spatial Map | Feature Reduction | AUROC (95% CI) | AUPRC (95% CI) | Accuracy (95% CI) | Sensitivity (95% CI) | Specificity (95% CI) |
| --- | --- | --- | --- | --- | --- | --- | --- |
| Transformer (L2 = 0) | Distance | DAE | **0.97 (0.90-1.00)** | **0.85 (0.60-1.00)** | **0.96 (0.90-1.00)** | **1.00 (1.00-1.00)** | 0.95 (0.87-1.00) |
| Transformer (D = 0.8) | Distance | DAE | **0.97 (0.90-1.00)** | 0.80 (0.52-1.00) | 0.95 (0.89-1.00) | **1.00 (0.94-1.00)** | 0.93 (0.86-1.00) |
| Transformer (D = 0) | Distance | DAE | 0.95 (0.89-1.00) | 0.77 (0.55-1.00) | **0.96 (0.91-1.00)** | **1.00 (1.00-1.00)** | 0.95 (0.88-1.00) |
| Transformer | Distance | DAE | 0.95 (0.90-0.99) | **0.85 (0.66-0.99)** | 0.91 (0.87-0.98) | **1.00 (0.90-1.00)** | 0.88 (0.80-0.99) |
| Transformer (L2 = 1e-2) | Distance | DAE | 0.91 (0.81-0.98) | 0.74 (0.51-0.96) | 0.91 (0.84-0.98) | 0.81 (0.70-1.00) | 0.95 (0.84-1.00) |
| Transformer (L2 = 1e-6) | Distance | DAE | 0.90 (0.77-0.99) | **0.85 (0.63-0.98)** | 0.93 (0.81-0.98) | 0.81 (0.71-1.00) | **0.97 (0.77-1.00)** |
| Transformer | None | DAE | 0.86 (0.66-0.99) | **0.85 (0.66-0.97)** | 0.88 (0.81-0.96) | 0.88 (0.63-1.00) | 0.88 (0.80-1.00) |
| Transformer (D = 0.2) | Distance | DAE | 0.83 (0.67-0.98) | 0.70 (0.49-0.95) | 0.93 (0.88-0.98) | 0.88 (0.70-1.00) | 0.95 (0.88-1.00) |
| XGBoost | None | None | 0.81 (0.68-0.93) | 0.65 (0.40-0.92) | 0.80 (0.59-0.95) | 0.62 (0.46-1.00) | 0.88 (0.47-1.00) |
| XGBoost | Coordinates | None | 0.78 (0.64-0.89) | 0.61 (0.35-0.82) | 0.77 (0.57-0.89) | 0.69 (0.44-1.00) | 0.80 (0.37-0.97) |
| Transformer | Distance | None | 0.76 (0.59-0.92) | 0.49 (0.29-0.74) | 0.84 (0.73-0.91) | 0.81 (0.66-1.00) | 0.85 (0.68-0.94) |
| RNN | Coordinates | None | 0.70 (0.56-0.83) | 0.39 (0.23-0.65) | 0.73 (0.58-0.83) | 0.69 (0.57-1.00) | 0.75 (0.40-0.88) |
| Transformer (L2 = 1e-4) | Distance | DAE | 0.65 (0.50-0.78) | 0.38 (0.16-0.66) | 0.50 (0.41-0.77) | **1.00 (0.42-1.00)** | 0.30 (0.20-0.88) |
| RNN | None | DAE | 0.63 (0.52-0.76) | 0.32 (0.22-0.49) | 0.62 (0.52-0.75) | 0.94 (0.81-1.00) | 0.50 (0.33-0.68) |
| RNN | Coordinates | DAE | 0.63 (0.50-0.79) | 0.32 (0.19-0.49) | 0.62 (0.47-0.76) | 0.94 (0.81-1.00) | 0.50 (0.32-0.68) |
| RNN | None | None | 0.61 (0.44-0.81) | 0.38 (0.22-0.58) | 0.77 (0.62-0.88) | 0.44 (0.28-0.82) | 0.90 (0.59-0.97) |
| Transformer (LR = 1e-3) | Distance | DAE | 0.57 (0.39-0.73) | 0.38 (0.21-0.61) | 0.55 (0.45-0.84) | 0.75 (0.18-0.97) | 0.47 (0.28-1.00) |
| Logistic | Coordinates | None | 0.54 (0.36-0.71) | 0.30 (0.17-0.54) | 0.50 (0.31-0.77) | 0.81 (0.18-1.00) | 0.38 (0.06-0.96) |
| Logistic | None | None | 0.52 (0.36-0.66) | 0.29 (0.15-0.49) | 0.46 (0.28-0.77) | 0.81 (0.20-1.00) | 0.33 (0.08-0.95) |

DL models were trained with an Adam optimizer [60], a default learning rate of 1e-2, weight decay of 1e-4, a batch size of 12 patients, 20% dropout rate, and standard binary cross-entropy loss. Because there is no logical order to the segmented glomeruli from our panoptic segmentation algorithm, when training the RNN baseline we shuffled the order of each WSI glomeruli set and each cross-validation iteration. All DL models used 20% of the training samples as a validation set; training was halted if loss on the validation set did not decrease after 20 epochs. A learning rate scheduler was used, which automatically decreased the learning rate if the validation loss did not improve after 10 epochs.

## 3. DATA

### 3.1 Data Acquisition

We utilized an existing cohort of 56 patients with type 2 diabetes characterized in greater detail in our prior work studying the integration of histology with urinary proteomics [53]. Study subjects were from Seoul National University Hospital. Human data collection followed protocols approved by the Institutional Review Board at the Seoul National University (SNU) College of Medicine (H-1812-159-998), Seoul, Korea. All experiments were performed according to federal guidelines and regulations. Patient data included demographics, medical history, kidney biopsy, and blood tests measured at the time of biopsy. Serum creatinine was used to determine estimated glomerular filtration rate (eGFR). Additional creatinine data was collected one year and two years following initial kidney biopsy, and eGFR was used to determine the presence of ESRD. In this work, we focus on the prediction of longer-term ESRD (measured two years after initial biopsy) from WSI data extracted at the time of initial biopsy. Biopsy WSIs were stained using a PAS procedure. An example biopsy from our dataset is shown in **Figure 3**.

**Figure 3.**
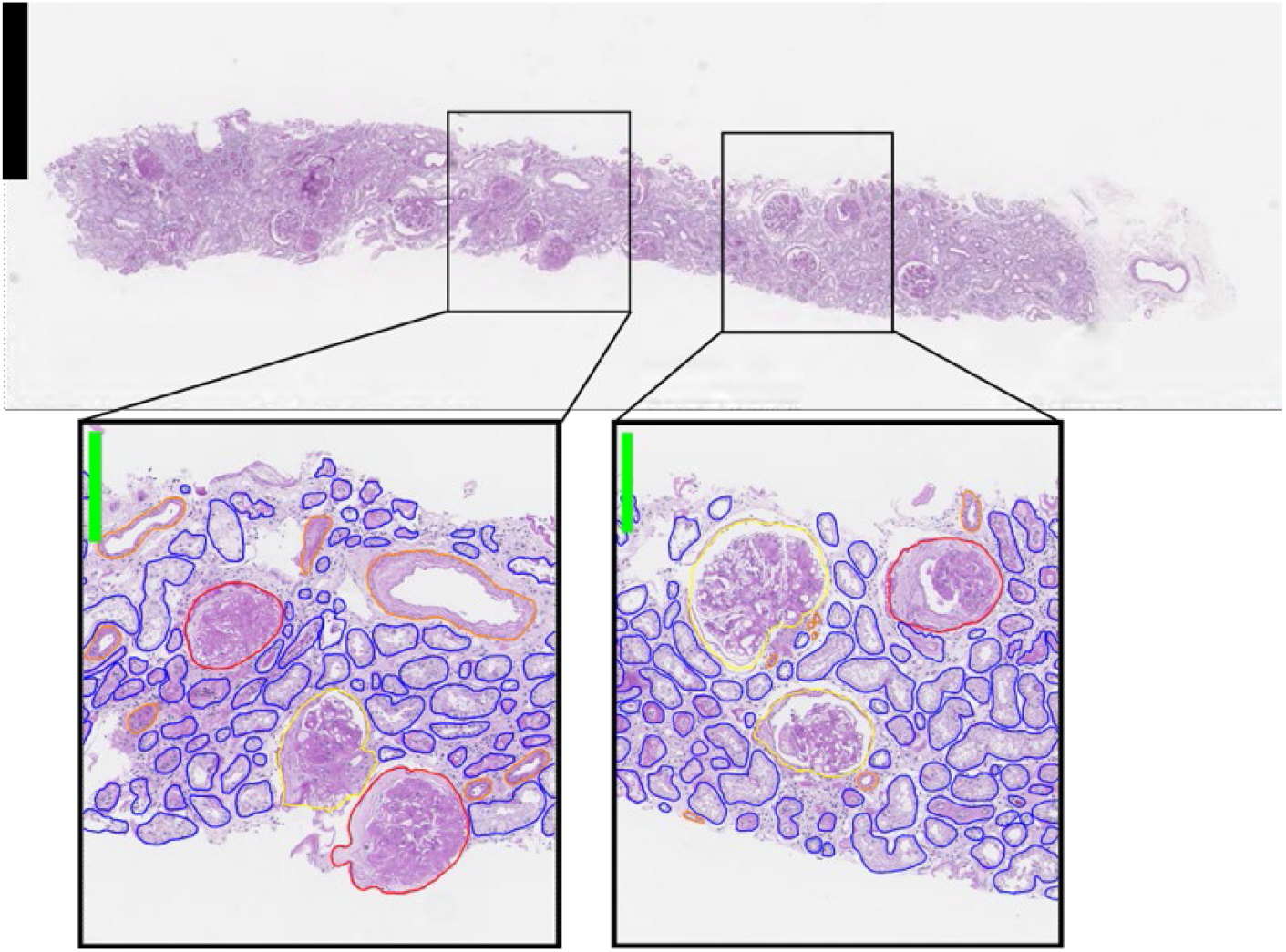
Whole slide image from our dataset corresponding to a patient with diabetic nephropathy whose two-year clinical follow-up confirmed end-stage renal disease. Select glomeruli detected by our pre-trained panoptic segmentation algorithm are shown in the lower region, along with other structures not considered for this study. Estimated centroid coordinates of each glomerulus are used to compute pairwise Euclidean distances and derive a learned relative positional embedding that is integrated into the transformer’s self-attention equation.

### 3.2 Data Processing

Panoptic segmentation was performed on PAS-stained WSIs to classify pixels into 6 categories, and to resolve instances of the same class. These 6 categories include image background, renal interstitium, nonglobally sclerotic glomeruli, globally sclerotic glomeruli, renal tubules, and arteries/arterioles. Medullary regions were manually annotated and excluded from analysis. The panoptic network was trained on previously annotated WSIs, using our previously published, publicly available H-AI-L pipeline [61]. Using the WSI panoptic classifications, glomerular and tubular pixels were further classified into three components based on their appearance in the PAS-stained biopsies: (1) nuclear components, (2) PAS-positive components, and (3) white space/luminal components. Nuclei were segmented using another separately trained panoptic classifier. The PAS+ and luminal components were then segmented using image thresholding.

Following the segmentation step, we extracted 316 digital image feature types (e.g., sclerosis, color, texture, morphology, containment, and inter/intrastructural distances) for each segmented glomerulus as detailed in prior work [53]. All features were independently standardized using z-normalization (using only values from the training set of each cross-validation iteration) to yield a per-feature distribution with a mean of zero and standard deviation of one.

Each patient in the dataset was associated with a single WSI. Each WSI contained a variable number of segmented glomeruli (mean: 16.6, 25^th^ percentile: 11.5, 50^th^: 15, 75^th^: 21.3, minimum: 2, maximum: 45).

## 4. RESULTS

We obtained a two-year ESRD probability for each of our 56 patients following a leave-one-out cross-validation procedure described in Section 2.5. Optimal classification thresholds were obtained from the Youden index [62]. Full classification results for each model variation are shown in **Table 1**. For brevity, we only list hyperparameter changes for the transformer and RNN that deviated from our baseline setting. Our baseline setting was 0.5 dropout, 0.0001 L2 weight regularization, and a learning rate of 0.01.

Our transformer network with no regularization, DAE-encoded features, and spatial pairwise distance embedding was the most accurate ESRD prediction model, achieving an AUROC of 0.97 (95% CI: 0.90-1.00), AUPRC of 0.85 (95% CI: 0.60-1.00), accuracy of 0.96 (95% CI:0.90-1.00), sensitivity of 1.00 (95% CI: 1.00-1.00), and a specificity of 0.95 (95% CI: 0.87-1.00).

In contrast, a similar transformer model *without* spatial awareness (i.e., no distance-based positional embedding) performed acceptably but measurably worse across most metrics, achieving an AUROC of 0.86 (95% CI: 0.66-0.99), AUPRC of 0.85 (0.66-0.97), accuracy of 0.88 (0.81-0.96), sensitivity of 0.88 (0.63-1.00), and a specificity of 0.88 (0.80-1.00). The transformer that did not use DAE for data compression and dimensionality reduction performed poorly, achieving an AUROC of 0.76 (95% CI:0.59-0.92), AUPRC of 0.49 (95% CI:0.29-0.74), accuracy of 0.84 (0.73-0l.91), sensitivity of 0.81 (0.66-1.00), and specificity of 0.85 (95% CI: 0.68-0.94).

Given its speed and simplicity, and coarse feature aggregation, the XGBoost baseline classifier performed reasonably well, yielding an AUROC of 0.81 (95% CI: 0.68-0.93), AUPRC of 0.65 (0.40-0.92), accuracy of 0.80 (95% CI:0.59-0.95), sensitivity of 0.62 (95% CI: 0.46-1.00), and specificity of 0.88 (0.47-1.00). Logistic regression performed poorly and was not able to learn from the extracted features. DAE encodings did not improve baseline performance and are withheld from **Table 1** for succinctness.

## 5. CONCLUSIONS

This initial work demonstrates the feasibility of a novel multi-stage pipeline and risk estimation framework for predicting ESRD two years before official diagnosis in patients with diabetic nephropathy in the setting of type 2 diabetes. Our approach, limited to glomeruli information available in whole slide histology images, was able to predict two-year ESRD with high accuracy, suggesting the likelihood of important contextual information contained in spatial histological interactions.

Our overall framework integrates convolutional neural networks (panoptic glomeruli segmentation), expert defined knowledge (sclerosis, color, texture, and morphological histology features), nonlinear transformation and dimensionality reduction (DAE), and transformer-based self-attention networks with a novel implementation of a pairwise distance embedding based on Euclidean distance between glomeruli centroids. Our framework was able to aggregate a variable-length set of extracted glomeruli features into a fixed-dimensional WSI context that could be used to accurately predict two-year ESRD.

The classification results in **Table 1** demonstrate the power of our spatial embedding mechanism for deriving a salient WSI context in pooling the individual glomerulus instance representations for each patient (an AUROC of 0.97 with spatial distance information; AUROC of 0.86 without). Additionally, our results suggest that handcrafted image features should be paired with a dimensionality reduction approach such as an autoencoder (AUROC of 0.97 with DAE; AUROC of 0.76 without). While the wide confidence intervals exemplify the unavoidable variability and challenges of applying DL to a small dataset, our approach presents several intentional decisions (handcrafted features, dimensionality reduction, leave-one-out cross-validation, experiments with regularization and dropout) that demonstrate it is not a futile endeavor.

This preliminary work holds value as a component of a clinical decision support system for diabetic nephropathy patients with type 2 diabetes. A digital tool that can accurately predict two-year ESRD can better inform pathologists and caregivers about potential risk and, with adequate validation and explainability, can influence therapies and lead to personalized treatment strategies in anticipation of suspected future ESRD. Additionally, pathology is a heterogenous field in which different pathologists may disagree in their interpretation or estimation. An AI tool can be used to provide quantitative assessment to digital pathology images that can offer supporting evidence or insights to complement existing pathology and clinical workflows.

This study is limited to data from a single institution. While classification metrics are high, they may be in part due to high variability and artifacts of working with a very small dataset (n=56); as such, we do not attempt to claim wider generalizability of our results, but present and justify our work as a pilot study for future investigation and highlight the relative differences between models’ design choices (e.g., spatial awareness, feature compression).

In future work, we will investigate additional graph-inspired methods of structural histology representation, explore spatial adaptations of vision transformers with raw gigapixel images, integrate multimodal data sources like patient demographics and electronic health record (EHR) data, and visualize self-attention weights to better understand spatial glomeruli connections and relative contributions for patient-level prediction from WSIs.

## Data Availability

All data produced in the present study are available upon reasonable request to the authors

## Acknowledgments

The project was supported by NIDDK grant R01 DK114485 (PS), a glue grant (PS) from the NIDDK Kidney Precision Medicine Project, a multi-disciplinary small team grant RSG201047.2 (PS) from the State University of New York, a pilot grant (PS) from the University of Buffalo’s Clinical and Translational Science Institute (CTSI) grant 3UL1TR00141206 S1, a DiaComp Pilot & Feasibility Project 21AU4180 (PS) with support from NIDDK Diabetic Complications Consortium grants U24 DK076169 and U24 DK115255, NIH-OD grant U54 HL145608 (PS), and NIDDK grant R01 DK131189 (PS).

